# Colchicine in patients admitted to hospital with COVID-19 (RECOVERY): a randomised, controlled, open-label, platform trial

**DOI:** 10.1101/2021.05.18.21257267

**Authors:** RECOVERY Collaborative Group, Peter W Horby, Mark Campbell, Enti Spata, Jonathan R Emberson, Natalie Staplin, Guilherme Pessoa-Amorim, Leon Peto, Martin Wiselka, Laura Wiffen, Simon Tiberi, Ben Caplin, Caroline Wroe, Christopher Green, Paul Hine, Benjamin Prudon, Tina George, Andrew Wight, J Kenneth Baillie, Buddha Basnyat, Maya H Buch, Lucy C Chappell, Jeremy N Day, Saul N Faust, Raph L Hamers, Thomas Jaki, Edmund Juszczak, Katie Jeffery, Wei Shen Lim, Alan Montgomery, Andrew Mumford, Kathryn Rowan, Guy Thwaites, Marion Mafham, Richard Haynes, Martin J Landray

## Abstract

**Background:** Colchicine has been proposed as a treatment for COVID-19 on the basis of its anti-inflammatory actions.

**Methods:** In this randomised, controlled, open-label trial, several possible treatments were compared with usual care in patients hospitalised with COVID-19. Eligible and consenting adults were randomly allocated in a 1:1 ratio to either usual standard of care alone or usual standard of care plus colchicine twice daily for 10 days or until discharge (or one of the other treatment arms) using web-based simple (unstratified) randomisation with allocation concealment. The primary outcome was 28-day mortality. The trial is registered with ISRCTN (50189673) and clinicaltrials.gov (NCT04381936).

**Findings:** Between 27 November 2020 and 4 March 2021, 5610 patients were randomly allocated to receive colchicine and 5730 patients to receive usual care alone. Overall, 1173 (21%) patients allocated to colchicine and 1190 (21%) patients allocated to usual care died within 28 days (rate ratio 1.01; 95% confidence interval [CI] 0.93-1.10; p=0.77). Consistent results were seen in all pre-specified subgroups of patients. There was no significant difference in duration of hospitalisation (median 10 days vs. 10 days) or the proportion of patients discharged from hospital alive within 28 days (70% vs. 70%; rate ratio 0.98; 95% CI 0.94-1.03; p=0.44). Among those not on invasive mechanical ventilation at baseline, there was no significant difference in the proportion meeting the composite endpoint of invasive mechanical ventilation or death (25% vs. 25%; risk ratio 1.02; 95% CI 0.96-1.09; p=0.47).

**Interpretation:** In adults hospitalised with COVID-19, colchicine was not associated with reductions in 28-day mortality, duration of hospital stay, or risk of progressing to invasive mechanical ventilation or death.

**Funding:** UK Research and Innovation (Medical Research Council) and National Institute of Health Research (Grant ref: MC_PC_19056). Wellcome Trust (Grant Ref: 222406/Z/20/Z) through the COVID-19 Therapeutics Accelerator.

## INTRODUCTION

Inflammation is a key feature of severe COVID-19. Markedly raised levels of inflammatory markers such as C-reactive protein (CRP), ferritin, interleukin-6 (IL-6) and other cytokines are observed in severe cases and are associated with poor outcomes.^1-5^ Inflammation is particularly prominent in the lung and vascular endothelium, and is commonly associated with extensive alveolar damage and thrombosis of large and small pulmonary vessels.^6^ Corticosteroids and interleukin-6 inhibitors have both been shown to reduce mortality in patients with severe COVID-19, while Janus kinase (JAK) inhibitors accelerate improvement in clinical status.^7-10^ Together these results show that inflammation is modifiable and anti-inflammatory therapy can improve clinical outcomes.

Inflammasomes are a key part of the innate immune response to SARS-CoV-2 infection. These cytosolic pattern recognition receptor systems are activated in response to detection of pathogens in the cytosol and stimulate the release of proinflammatory cytokines.^11^ In COVID-19, the degree of inflammasome activation, particularly the nucleotide binding domain (NOD)-like pyrin domain 3 (NLRP3) inflammasome, correlates with disease severity.^12^ Colchicine, a readily available, safe and inexpensive drug, has a wide range of anti-inflammatory effects, including inhibition of the NLRP3 inflammasome.^13^ In addition to its role in treating acute gout and pericarditis, there is emerging evidence that colchicine may inhibit endovascular inflammation and provide clinical benefits in patients with coronary artery disease.^14-17^ Given the activation of NLRP3 in COVID-19 and the presence of vascular endothelial inflammation, colchicine has been proposed as a treatment for SARS-CoV-2 associated inflammatory disease.

However, only three small randomised controlled trials have assessed the effects of colchicine in hospitalised patients and, with a total of only seven deaths across these studies combined, none were adequately powered to identify any impact on mortality.^18-20^ Here we report the results of a large randomised controlled trial of colchicine in patients hospitalised with COVID-19.

## METHODS

### Study design and participants

The Randomised Evaluation of COVID-19 therapy (RECOVERY) trial is an investigator-initiated, individually randomised, controlled, open-label, platform trial to evaluate the effects of potential treatments in patients hospitalised with COVID-19. Details of the trial design and results for other possible treatments (dexamethasone, hydroxychloroquine, lopinavir-ritonavir, azithromycin, tocilizumab, and convalescent plasma) have been published previously.^7,9,21-24^ The trial is underway at 177 hospitals in the United Kingdom supported by the National Institute for Health Research Clinical Research Network, two hospitals in Indonesia, and two hospitals in Nepal (appendix pp 3-25). The trial is coordinated by the Nuffield Department of Population Health at University of Oxford (Oxford, UK), the trial sponsor. The trial is conducted in accordance with the principles of the International Conference on Harmonisation–Good Clinical Practice guidelines and approved by the UK Medicines and Healthcare products Regulatory Agency (MHRA) and the Cambridge East Research Ethics Committee (ref: 20/EE/0101). The protocol, statistical analysis plan, and additional information are available on the study website www.recoverytrial.net.

Patients admitted to hospital were eligible for the study if they had clinically suspected or laboratory confirmed SARS-CoV-2 infection and no medical history that might, in the opinion of the attending clinician, put the patient at significant risk if they were to participate in the trial. Children and pregnant women were not eligible for randomisation to colchicine. Patients with severe liver impairment, significant cytopaenia, concomitant use of strong CYP3A4 or P-glycoprotein inhibitors, or hypersensitivity to lactose were excluded (further details in appendix p 80). Written informed consent was obtained from all patients, or a legal representative if patients were too unwell or unable to provide consent.

### Randomisation and masking

Baseline data were collected using a web-based case report form that included demographics, level of respiratory support, major comorbidities, suitability of the study treatment for a particular patient, and treatment availability at the study site (appendix pp 32-34). Eligible and consenting, non-pregnant adult patients were assigned in a 1:1 ratio to either usual standard of care or usual standard of care plus colchicine or one of the other available RECOVERY treatment arms using web-based simple (unstratified) randomisation with allocation concealed until after randomisation (appendix pp 30-31). For some patients, colchicine was unavailable at the hospital at the time of enrolment or was considered by the managing physician to be either definitely indicated or definitely contraindicated. These patients were excluded from the randomised comparison between colchicine and usual care. Patients allocated to colchicine were to receive 1 mg after randomisation followed by 500 mcg 12 hours later and then 500 mcg twice daily by mouth or nasogastric tube for 10 days in total or until discharge, whichever occurred earlier. Dose frequency was halved for patients receiving a moderate CYP3A4 inhibitor or who had renal impairment (estimated glomerular filtration rate <30 ml/min/1.73m^2^) or estimated body weight <70 kg (appendix p 80).

As a platform trial, and in a factorial design, patients could be simultaneously randomised to other treatment groups: i) convalescent plasma versus monoclonal antibody (REGN-CoV2) versus usual care, ii) aspirin versus usual care, and iii) baricitinib versus usual care (appendix pp 31). Until 24 January 2021, the trial also allowed a subsequent randomisation for patients with progressive COVID-19 (evidence of hypoxia and a hyper-inflammatory state) to tocilizumab versus usual care. Participants and local study staff were not masked to the allocated treatment. The trial steering committee, investigators, and all other individuals involved in the trial were masked to outcome data during the trial.

### Procedures

A single online follow-up form was completed when participants were discharged, had died or at 28 days after randomisation, whichever occurred earliest (appendix pp 35-41). Information was recorded on adherence to allocated study treatment, receipt of other COVID-19 treatments, duration of admission, receipt of respiratory or renal support, and vital status (including cause of death). In addition, in the UK, routine healthcare and registry data were obtained including information on vital status (with date and cause of death), discharge from hospital, receipt of respiratory support, or renal replacement therapy.

### Outcomes

Outcomes were assessed at 28 days after randomisation, with further analyses specified at 6 months. The primary outcome was all-cause mortality. Secondary outcomes were time to discharge from hospital, and, among patients not on invasive mechanical ventilation at randomisation, invasive mechanical ventilation (including extra-corporal membrane oxygenation) or death. Prespecified subsidiary clinical outcomes were use of non-invasive respiratory support, time to successful cessation of invasive mechanical ventilation (defined as cessation of invasive mechanical ventilation within, and survival to, 28 days), use of renal dialysis or haemofiltration, cause-specific mortality, bleeding events, thrombotic events, and major cardiac arrhythmias. Information on suspected serious adverse reactions was collected in an expedited fashion to comply with regulatory requirements.

### Statistical Analysis

The primary analysis for all outcomes was by intention-to-treat comparing patients randomised to colchicine with patients randomised to usual care but for whom colchicine was both available and suitable as a treatment. For the primary outcome of 28-day mortality, the log-rank observed minus expected statistic and its variance were used to both test the null hypothesis of equal survival curves (i.e., the log-rank test) and to calculate the one-step estimate of the average mortality rate ratio. We constructed Kaplan-Meier survival curves to display cumulative mortality over the 28-day period. We used the same method to analyse time to hospital discharge and successful cessation of invasive mechanical ventilation, with patients who died in hospital right-censored on day 29. Median time to discharge was derived from Kaplan-Meier estimates. For the pre-specified composite secondary outcome of progression to invasive mechanical ventilation or death within 28 days (among those not receiving invasive mechanical ventilation at randomisation), and the subsidiary clinical outcomes of receipt of ventilation and use of haemodialysis or haemofiltration, the precise dates were not available and so the risk ratio was estimated instead.

Prespecified analyses were performed for the primary outcome using the statistical test of interaction (test for heterogeneity or trend), in accordance with the prespecified analysis plan, defined by characteristics at randomisation: age, sex, ethnicity, level of respiratory support, days since symptom onset, and use of corticosteroids (appendix p 114).

Estimates of rate and risk ratios are shown with 95% confidence intervals. All p-values are 2-sided and are shown without adjustment for multiple testing. The full database is held by the study team which collected the data from study sites and performed the analyses at the Nuffield Department of Population Health, University of Oxford (Oxford, UK).

As stated in the protocol, appropriate sample sizes could not be estimated when the trial was being planned at the start of the COVID-19 pandemic (appendix p 54). As the trial progressed, the trial steering committee, whose members were unaware of the results of the trial comparisons, determined that sufficient patients should be enrolled to provide at least 90% power at a two-sided significance level of 0.01 to detect a clinically relevant proportional reduction in 28-day mortality of 12.5% between the two groups.

On 4 March 2021, the independent data monitoring committee (DMC) conducted a routine review of the available safety and efficacy data. The DMC notified the chief investigators that there was no convincing evidence that further recruitment to the colchicine comparison would provide conclusive proof of worthwhile mortality benefit either overall or in any pre-specified subgroup. Consequently, recruitment to the colchicine comparison was closed on 5 March 2021 and preliminary results were made available to the public.

Analyses were performed using SAS version 9.4 and R version 3.4. The trial is registered with ISRCTN (50189673) and clinicaltrials.gov (NCT04381936).

### Role of the funding source

The funder of the study had no role in study design, data collection, data analysis, data interpretation, or writing of the report. The corresponding authors had full access to all the data in the study and had final responsibility for the decision to submit for publication.

## RESULTS

Between 27 November 2020 and 4 March 2021, 11,340 (58%) of 19423 patients enrolled into the RECOVERY trial were eligible to be randomly allocated to colchicine (i.e. colchicine was available in the hospital at the time and the attending clinician was of the opinion that the patient had no known indication for or contraindication to colchicine, figure 1). 5610 patients were randomly allocated to colchicine and 5730 were randomly allocated to usual care (36 patients were randomised outside of UK). The mean age of study participants in this comparison was 63.4 years (SD 13.8) and the median time since symptom onset was 9 days (IQR 6 to 12 days) (webtable 1). At randomisation, 10603 (94%) of patients were receiving corticosteroids.

**Figure 1:**
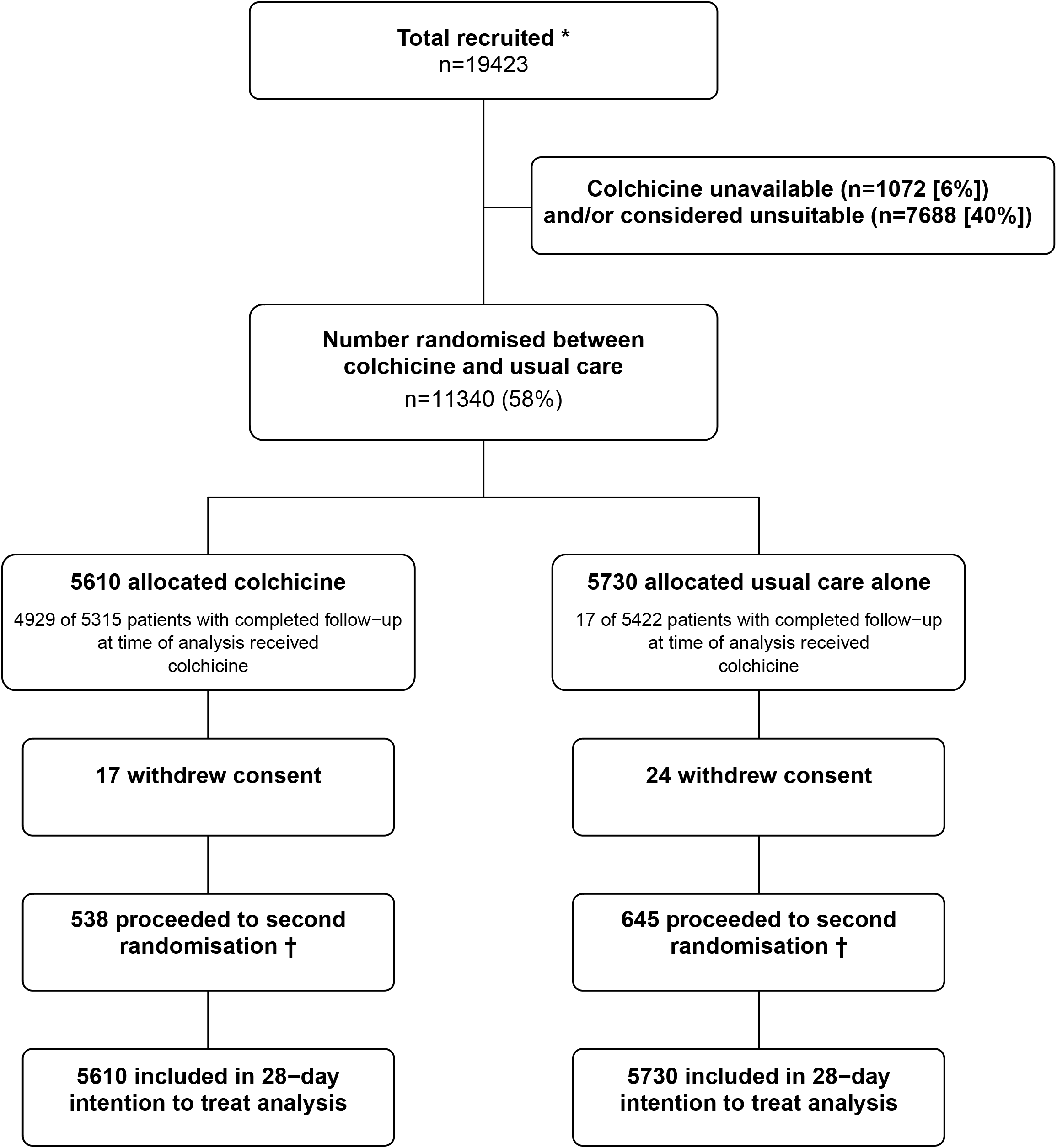
Trial profile. ITT=intention to treat. * Number recruited overall during period that adult participants could be recruited into colchicine comparison. Of the 11,340 randomised to colchicine vs usual care, 7091 were additionally randomised to convalescent plasma vs REGN−COV2 vs usual care (3505 [62%] of the colchicine group vs 3586 [63%] of the usual care group); 7545 were additionally randomised to aspirin vs usual care (3747 [67%] of the colchicine group vs 3798 [66%] of the usual care group), and 1635 patients were additionally randomised to baricitinib vs usual care (802 [14%] of the colchicine group vs 833 [15%] of the usual care group). † Includes 251/5610 (4%) patients in the colchicine arm and 306/5730 (5%) patients in the usual care arm allocated to tocilizumab.

The follow-up form was completed for 5510 (98%) in the colchicine group and 5605 (98%) in the usual care group. Among patients with a completed follow-up form, 5122 (93%) allocated to colchicine received at least one dose (figure 1; webtable 2). The median duration of treatment with colchicine was 6 days (IQR 3-9 days). Use of other treatments for COVID-19 was similar among patients allocated colchicine and among those allocated usual care, with 87% receiving a corticosteroid, about one-quarter receiving remdesivir, and one-eighth receiving tocilizumab (webtable 2).

Primary and secondary outcome data are known for >99% of randomly assigned patients. There was no significant difference in the proportion of patients who met the primary outcome of 28-day mortality between the two randomised groups (1173 [21%] patients in the colchicine group vs. 1190 (21%) patients in the usual care group; rate ratio 1.01; 95% confidence interval [CI], 0.93 to 1.10; p=0.77; figure 2). We observed similar results across all pre-specified sub-groups (figure 3). In an exploratory analysis restricted to the 11009 (97%) patients with a positive SARS-CoV-2 test result, the result was virtually identical (rate ratio 1.02, 95% CI 0.94 to 1.10; p=0.70).

**Figure 2:**
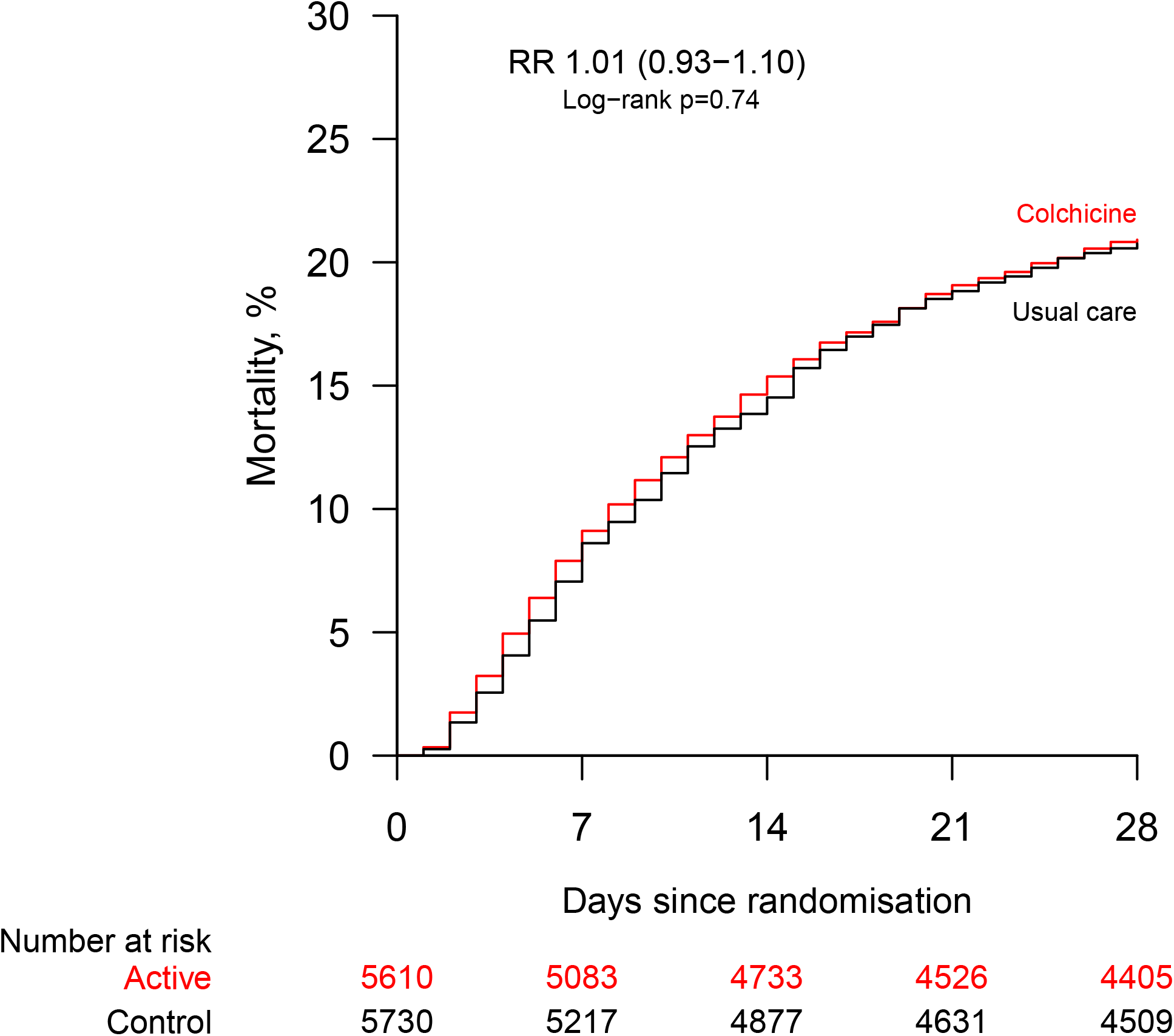
Effect of allocation to colchicine on 28-day mortality.

**Figure 3:**
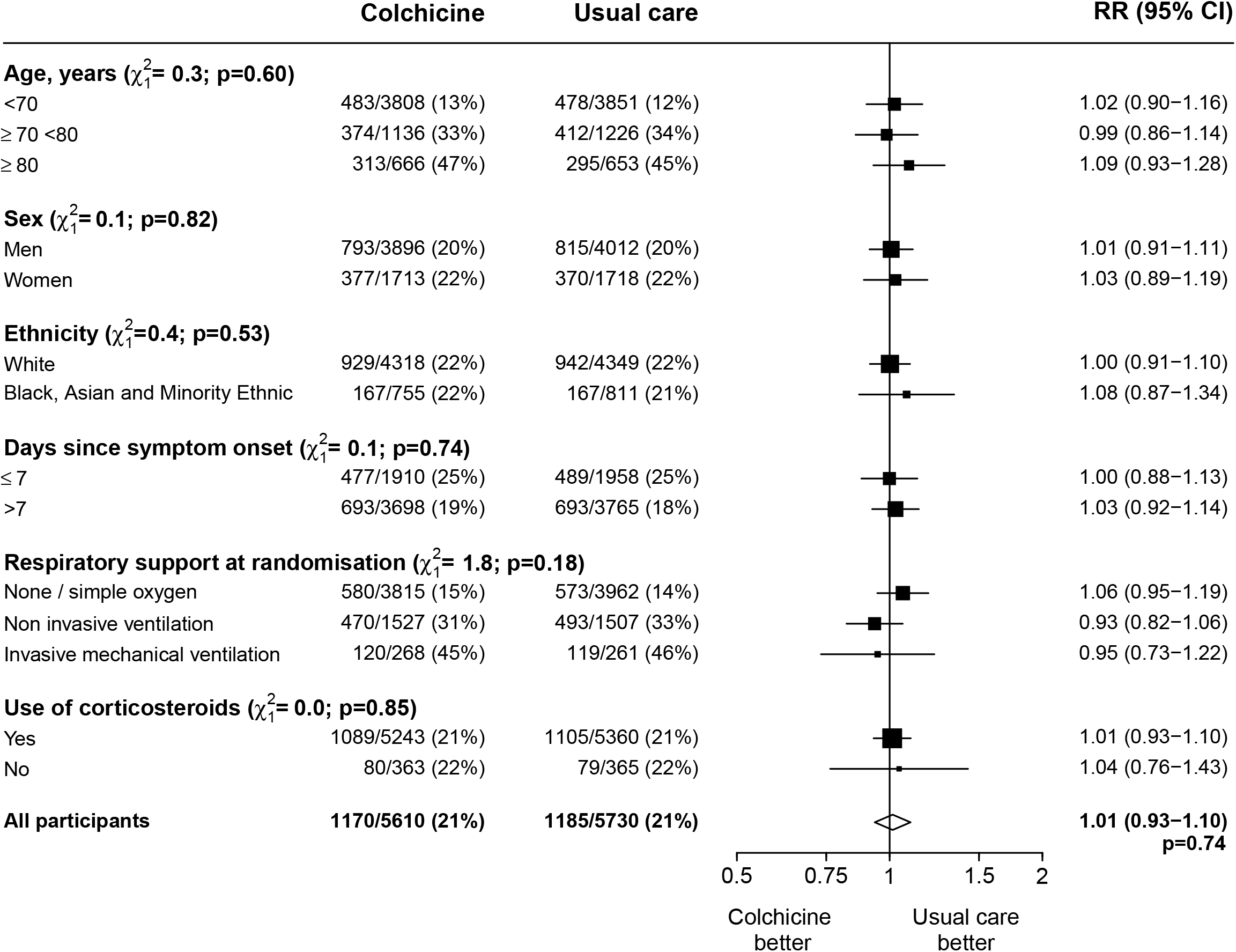
Effect of allocation to colchicine on 28-day mortality by baseline characteristics. Subgroup−specific rate ratio estimates are represented by squares (with areas of the squares proportional to the amount of statistical information) and the lines through them correspond to the 95% CIs. The ethnicity, days since onset and use of corticosteroids subgroups exclude those with missing data, but these patients are included in the overall summary diamond.

The median time to discharge from hospital alive was 10 days (IQR 5 to >28) in both groups and there was no significant difference in the probability of being discharged alive within 28 days (70% vs. 70%, rate ratio 0.98, 95% CI 0.94 to 1.03, p=0.44) (table 2). Among those not on invasive mechanical ventilation at baseline, the number of patients progressing to the pre-specified composite secondary outcome of invasive mechanical ventilation or death was similar in both groups (25% vs. 25%, risk ratio 1.02, 95% CI 0.96 to 1.09, p=0.47). Similar results were seen in all pre-specified subgroups of patients (webfigure 1, webfigure 2).

**Table 1:** Baseline characteristics.

|  | Treatment allocation |  |
| --- | --- | --- |
|  | Colchicine<br>(n=5610) | Usual care<br>(n=5730) |
| Age, years | 63.3 (13.8) | 63.5 (13.7) |
| <70 | 3806 (68%) | 3850 (67%) |
| ≥70 to <80 | 1139 (20%) | 1227 (21%) |
| ≥80 | 665 (12%) | 653 (11%) |
| Sex |  |  |
| Male | 3896 (69%) | 4012 (70%) |
| Female | 1713 (31%) | 1718 (30%) |
| Ethnicity |  |  |
| White | 4344 (77%) | 4383 (76%) |
| Black, Asian, and minority ethnic | 758 (14%) | 813 (14%) |
| Unknown | 508 (9%) | 534 (9%) |
| Number of days since symptom onset | 9 (6-12) | 9 (6-12) |
| Number of days since admission to hospital | 2 (1-3) | 2 (1-3) |
| Respiratory support received |  |  |
| None / simple oxygen | 3815 (68%) | 3962 (69%) |
| Non invasive ventilation | 1527 (27%) | 1507 (26%) |
| Invasive mechanical ventilation | 268 (5%) | 261 (5%) |
| Previous diseases |  |  |
| Diabetes | 1426 (25%) | 1470 (26%) |
| Heart disease | 1189 (21%) | 1231 (21%) |
| Chronic lung disease | 1208 (22%) | 1206 (21%) |
| Tuberculosis | 16 (<1%) | 13 (<1%) |
| HIV | 11 (<1%) | 20 (<1%) |
| Severe liver disease * | 0 (0%) | 0 (0%) |
| Severe kidney impairment † | 170 (3%) | 166 (3%) |
| Any of the above | 2880 (51%) | 2963 (52%) |
| Use of corticosteroids |  |  |
| Yes | 5243 (93%) | 5360 (94%) |
| No | 363 (6%) | 365 (6%) |
| Missing | 4 (<1%) | 5 (<1%) |
| Severe acute respiratory syndrome coronavirus 2 test result |  |  |
| Positive | 5456 (97%) | 5553 (97%) |
| Negative | 57 (1%) | 58 (1%) |
| Unknown | 97 (2%) | 119 (2%) |

Data are mean (SD), n (%), or median (IQR). No children or pregnant women were randomised. \* Defined as requiring ongoing specialist care. † Defined as estimated glomerular filtration rate <30 mL/min per 1.73 m<sup>2</sup>.

**Table 2:** Effect of allocation to colchicine on key study outcomes.

|  | Treatment allocation |  | RR (95% CI) | p-value |
| --- | --- | --- | --- | --- |
|  | Colchicine<br>(n=5610) | Usual care<br>(n=5730) |  |  |
| Primary outcome: |  |  |  |  |
| 28-day mortality | 1173 (21%) | 1190 (21%) | 1.01 (0.93-1.10) | 0.77 |
| Secondary outcomes: |  |  |  |  |
| Median time to being discharged alive, days | 10 (5 to >28) | 10 (5 to >28) |  |  |
| Discharged from hospital within 28 days | 3901 (70%) | 4032 (70%) | 0.98 (0.94-1.03) | 0.44 |
| Receipt of invasive mechanical ventilation or death* | 1344/5342 (25%) | 1343/5469 (25%) | 1.02 (0.96-1.09) | 0.47 |
| Invasive mechanical ventilation | 600/5342 (11%) | 591/5469 (11%) | 1.04 (0.93-1.16) | 0.48 |
| Death | 1053/5342 (20%) | 1070/5469 (20%) | 1.01 (0.93-1.09) | 0.85 |
| Subsidiary clinical outcomes |  |  |  |  |
| Receipt of ventilation † | 852/3815 (22%) | 941/3962 (24%) | 0.94 (0.87-1.02) | 0.14 |
| Non-invasive ventilation | 818/3815 (21%) | 904/3962 (23%) | 0.94 (0.86-1.02) | 0.14 |
| Invasive mechanical ventilation | 259/3815 (7%) | 228/3962 (6%) | 1.18 (0.99-1.40) | 0.06 |
| Successful cessation of invasive mechanical ventilation ‡ | 88/268 (33%) | 81/261 (31%) | 1.01 (0.75-1.37) | 0.93 |
| Use of haemodialysis or haemofiltration § | 212/5570 (4%) | 203/5683 (4%) | 1.07 (0.88-1.29) | 0.51 |

Data are n (%) or n/N (%), unless otherwise indicated. RR=rate ratio for the outcomes of 28-day mortality, hospital discharge and successful cessation of invasive mechanical ventilation, and risk ratio for other outcomes. \* Analyses exclude those on invasive mechanical ventilation at randomisation. † Analyses exclude those on any form of ventilation at randomisation. ‡ Analyses restricted to those on invasive mechanical ventilation at randomisation. § Analyses exclude those on haemodialysis or haemofiltration at randomisation.

We found no significant differences in the prespecified subsidiary clinical outcomes of cause-specific mortality (webtable 3), use of ventilation, successful cessation of invasive mechanical ventilation, or need for renal dialysis or haemofiltration (table 2). The incidence of new cardiac arrhythmias, bleeding events, and thrombotic events was also similar in the two groups (webtable 4). There were two reports of a serious adverse reaction believed related to colchicine: one case of severe acute kidney injury and one case of rhabdomyolysis.

## DISCUSSION

In this large, randomised trial involving over 11,000 patients from 3 countries and over 2000 deaths, allocation to colchicine was not associated with reductions in mortality, duration of hospitalisation or the risk of being ventilated or dying for those not on ventilation at baseline. These results were consistent across the prespecified subgroups of age, sex, ethnicity, duration of symptoms prior to randomisation, level of respiratory support at randomisation, and use of corticosteroids.

The benefit of dexamethasone in COVID-19 patients requiring respiratory support demonstrates the importance of inflammation in this patient group and colchicine was proposed as a treatment for COVID-19 based on its anti-inflammatory activity.^25^ The lack of evidence of benefit from colchicine in this large well-powered trial suggests that the anti-inflammatory properties of colchicine are either insufficient to produce a meaningful reduction in mortality risk or are not affecting the relevant inflammatory pathways in moderate to severe COVID-19. Whilst the majority of patients in this study received concomitant corticosteroid therapy, we saw no evidence that colchicine was beneficial in those patients not receiving a corticosteroid.

Strengths of this trial included that it was randomised, had a large sample size, broad eligibility criteria, was international and more than 99% of patients were followed up for the primary outcome. However, detailed information on laboratory markers of inflammation and immune response was not collected, nor was information on radiological or physiological outcomes. Although this randomised trial is open label (i.e., participants and local hospital staff are aware of the assigned treatment), the outcomes are unambiguous and were ascertained without bias through linkage to routine health records.

Three other randomised controlled trials have assessed the efficacy of colchicine for the treatment of COVID-19 in hospitalised patients.^18-20^ A two day shorter duration of hospitalisation was reported in a trial of 100 patients with laboratory confirmed SARS-CoV-2 infection and pulmonary involvement on computed tomography who were randomized to either hydroxychloroquine plus colchicine or hydroxychloroquine plus placebo.^18^ A second trial reported a reduced duration of hospitalisation and oxygen therapy in 36 in-patients allocated colchicine compared to 36 in-patients allocated usual care, which included hydroxychloroquine, azithromycin and methylprednisolone.^19^ Finally, the GRECCO-19 trial reported a lower rate of clinical deterioration in 55 patients randomly assigned to colchicine compared to 50 patients randomly assigned to usual care (which did not include corticosteroids).^20^ The total number of patients in all three prior trials combined was 285 with seven deaths during the follow-up period. By contrast, the RECOVERY trial, with more than 11,000 participants and more than 2,000 deaths, had excellent power to detect modest treatment benefits; none were observed.

The RECOVERY trial only studied patients who had been hospitalised with COVID-19 and, therefore, is not able to provide any evidence on the safety and efficacy of colchicine used in other patient groups. In the COLCORONA trial of 4488 non-hospitalised patients with laboratory confirmed or clinically suspected COVID-19, fewer patients in the colchicine group met the composite endpoint of death or hospitalisation within 30 days of randomization than in the placebo group. However, the trial was stopped before the scheduled sample size had been fully enrolled due to logistical reasons and the result was not statistically significant (odds ratio, 0.79; 95% CI 0.61 to 1.03; P=0.08).^26^ Thus the role of colchicine in treatment of COVID-19 among patients not requiring hospitalsation remains uncertain. Further trials in that setting are ongoing.^27^

In summary, the results of this large, randomised trial do not support the use of colchicine in adults hospitalised with COVID-19.

### Evidence before this study

We searched medRxiv, bioRxiv, Medline, Embase and the WHO International Clinical Trials Registry Platform from September 1, 2019 to April 1, 2021 for clinical trials evaluating the effect of colchicine treatment among patients hospitalised with COVID-19, using the search terms (“SARS-CoV-2.mp” OR “COVID.mp” OR “COVID-19.mp” OR “2019-nCoV.mp” OR “Coronavirus.mp” OR “Coronavirinae/”) AND (“colchicine.mp” OR “colchicine/”) in any language, using validated filters to select for randomised controlled trials.

We identified three relevant randomised trials that compared colchicine with usual care or placebo in hospitalised patients with COVID-19 (two at low risk of bias and one with some concerns due to limited information on randomisation process and lack of clarity about blinding in the study). Each trial suggested a potential favourable impact of colchicine on outcome measures of clinical improvement or duration of hospitalisation, The three trials combined included a total of 285 patients and 7 deaths, so even combined were not adequately powered to detect an effect on mortality.

### Added value of this study

The Randomised Evaluation of COVID-19 therapy (RECOVERY) trial is the first large, randomised trial to report results of the effect of colchicine in patients hospitalised with COVID-19. We found no significant effect of colchicine vs. usual care alone on 28-day mortality, the probability of discharge alive within 28 days, or, among patients who were not receiving invasive mechanical ventilation at randomisation, the probability of progressing to the composite outcome of invasive mechanical ventilation or death. We saw no evidence of benefit of colchicine in any patient subgroup.

### Implications of all the available evidence

Colchicine treatment is not of clinical benefit for adults hospitalised with COVID-19 compared with current usual care.

## Supporting information

CONSORT Checklist

## Data Availability

The protocol, consent form, statistical analysis plan, definition & derivation of clinical characteristics & outcomes, training materials, regulatory documents, and other relevant study materials are available online at www.recoverytrial.net. As described in the protocol, the trial Steering Committee will facilitate the use of the study data and approval will not be unreasonably withheld. Deidentified participant data will be made available to bona fide researchers registered with an appropriate institution within 3 months of publication. However, the Steering Committee will need to be satisfied that any proposed publication is of high quality, honours the commitments made to the study participants in the consent documentation and ethical approvals, and is compliant with relevant legal and regulatory requirements (e.g. relating to data protection and privacy). The Steering Committee will have the right to review and comment on any draft manuscripts prior to publication.
https://www.ndph.ox.ac.uk/data-access

## Contributors

This manuscript was initially drafted by the PWH and MJL, further developed by the Writing Committee, and approved by all members of the trial steering committee. PWH and MJL vouch for the data and analyses, and for the fidelity of this report to the study protocol and data analysis plan. PWH, JKB, MB, LCC, JD, SNF, TJ, EJ, KJ, WSL, AMo, AMu, KR, GT, MM, RH, and MJL designed the trial and study protocol. MM, MC, G P-A, LP, MW, LW, ST, BC, CW, CG, PH, BP, TG, AW, the Data Linkage team at the RECOVERY Coordinating Centre, and the Health Records and Local Clinical Centre staff listed in the appendix collected the data. ES, NS, and JRE did the statistical analysis. All authors contributed to data interpretation and critical review and revision of the manuscript. PWH and MJL had access to the study data and had final responsibility for the decision to submit for publication.

## Writing Committee (on behalf of the RECOVERY Collaborative Group)

Peter W Horby PhD FRCP, Mark Campbell FRCPath, Enti Spata, Jonathan R Emberson PhD, Natalie Staplin PhD, Guilherme Pessoa-Amorim, Leon Peto PhD, Martin Wiselka PhD, Laura Wiffen MRCP, Simon Tiberi FRCP, Ben Caplin PhD, Caroline Wroe PhD, Christopher Green DPhil, Paul Hine MRCP, Benjamin Prudon FRCP (Edin), Tina George MRCP, Andrew Wight MRCP, J Kenneth Baillie MD PhD, Buddha Basnyat, Maya Buch PhD FRCP, Lucy C Chappell PhD, Jeremy Day PhD FRCP, Saul N Faust FRCPCH, Raph L Hamers, Thomas Jaki PhD, Edmund Juszczak MSc, Katie Jeffery PhD, Wei Shen Lim FRCP, Alan Montgomery PhD, Andrew Mumford PhD, Kathryn Rowan PhD, Guy Thwaites PhD FRCP, Marion Mafham MD, Richard Haynes DM, Martin J Landray PhD FRCP.

## Data Monitoring Committee

Peter Sandercock, Janet Darbyshire, David DeMets, Robert Fowler, David Lalloo, Mohammed Munavvar (from January 2021), Ian Roberts (until December 2020), Janet Wittes.

## Declaration of interests

The authors have no conflict of interest or financial relationships relevant to the submitted work to disclose. No form of payment was given to anyone to produce the manuscript. All authors have completed and submitted the ICMJE Form for Disclosure of Potential Conflicts of Interest. The Nuffield Department of Population Health at the University of Oxford has a staff policy of not accepting honoraria or consultancy fees directly or indirectly from industry (see https://www.ndph.ox.ac.uk/files/about/ndph-independence-of-research-policy-jun-20.pdf).

## Data sharing

The protocol, consent form, statistical analysis plan, definition & derivation of clinical characteristics & outcomes, training materials, regulatory documents, and other relevant study materials are available online at www.recoverytrial.net. As described in the protocol, the trial Steering Committee will facilitate the use of the study data and approval will not be unreasonably withheld. Deidentified participant data will be made available to bona fide researchers registered with an appropriate institution within 3 months of publication. However, the Steering Committee will need to be satisfied that any proposed publication is of high quality, honours the commitments made to the study participants in the consent documentation and ethical approvals, and is compliant with relevant legal and regulatory requirements (e.g. relating to data protection and privacy). The Steering Committee will have the right to review and comment on any draft manuscripts prior to publication. Data will be made available in line with the policy and procedures described at: https://www.ndph.ox.ac.uk/data-access. Those wishing to request access should complete the form at https://www.ndph.ox.ac.uk/files/about/data_access_enquiry_form_13_6_2019.docx and e-mailed to

## Acknowledgements

Above all, we would like to thank the thousands of patients who participated in this trial. We would also like to thank the many doctors, nurses, pharmacists, other allied health professionals, and research administrators at 177 NHS hospital organisations across the whole of the UK, supported by staff at the National Institute of Health Research (NIHR) Clinical Research Network, NHS DigiTrials, Public Health England, Department of Health & Social Care, the Intensive Care National Audit & Research Centre, Public Health Scotland, National Records Service of Scotland, the Secure Anonymised Information Linkage (SAIL) at University of Swansea, and the NHS in England, Scotland, Wales and Northern Ireland.

The RECOVERY trial is supported by grants to the University of Oxford from UK Research and Innovation (UKRI) and NIHR (MC_PC_19056), the Wellcome Trust (Grant Ref: 222406/Z/20/Z) through the COVID-19 Therapeutics Accelerator, and by core funding provided by the NIHR Oxford Biomedical Research Centre, the Wellcome Trust, the Bill and Melinda Gates Foundation, the Foreign, Commonwealth and Development Office, Health Data Research UK, the Medical Research Council Population Health Research Unit, the NIHR Health Protection Unit in Emerging and Zoonotic Infections, and NIHR Clinical Trials Unit Support Funding. TJ is supported by a grant from UK Medical Research Council (MC_UU_0002/14) and an NIHR Senior Research Fellowship (NIHR-SRF-2015-08-001). WSL is supported by core funding provided by NIHR Nottingham Biomedical Research Centre. Combiphar supplied colchicine free of charge for use in this trial in Indonesia. Tocilizumab was provided free of charge for this trial by Roche Products Limited. REGN-COV2 was provided free of charge for this trial by Regeneron. Convalescent plasma was collected by NHS Blood and Transplant, the Scottish National Blood Transfusion Service, Welsh Blood Service, Northern Ireland Blood Transfusion Service and funded by the DHSC through core funding and funding under COVID-19 and EU SoHo Grant.

The views expressed in this publication are those of the authors and not necessarily those of the NHS, the NIHR, or the UK Department of Health and Social Care.

## Notes

### Competing Interest Statement

The authors have declared no competing interest.

### Clinical Trial

The trial is registered with ISRCTN (50189673) and clinicaltrials.gov (NCT04381936).

### Clinical Protocols

https://www.recoverytrial.net

### Author Declarations

The trial is conducted in accordance with the principles of the International Conference on Harmonisation-Good Clinical Practice guidelines and approved by the UK Medicines and Healthcare products Regulatory Agency (MHRA) and the Cambridge East Research Ethics Committee (ref: 20/EE/0101).

